# Qualitative exploration of patients’ experiences with Intrabeam targeted intraoperative radiotherapy (TARGIT-IORT) and External-Beam Radiotherapy Treatment (EBRT)

**DOI:** 10.1101/2023.09.14.23295478

**Authors:** Sandeep Bagga, Natalie Swiderska, Charlotte Hooker, Jenny Royle, Marie Ennis O’Connor, Siobhan Freeney, Dympna Watson, Robin Woolcock

**Affiliations:** MediPaCe; Independent Patient Advocate; Lobular (Breast Cancer) Ireland; UK Charity for Triple Negative Breast Cancer

**Keywords:** Patient experience, breast cancer, radiotherapy, qualitative methods, TARGIT-IORT, breast conserving surgery

## Abstract

For patients with early breast cancer undergoing breast conserving surgery, radiotherapy is given either as a post-operative course of External-Beam RadioTherapy (EBRT), given daily for over a number of days and often weeks, or with TARGeted Intrabeam Intraoperative radioTherapy (TARGIT-IORT), which is delivered, during surgery under the same anaesthetic. Several studies have reported the quantitative Quality-of-Life (QoL) benefits of TARGIT-IORT over EBRT. This qualitative study was designed to gather a deep understanding of the benefits and impacts of receiving EBRT or TARGIT-IORT as perceived by patients. It also captured how differently the treatments affected the lived experience of the patient and their care partner through their treatment journey.

A patient-led Working Group was established to guide study design, delivery and to help validate findings. Patients with experience of receiving EBRT or TARGIT-IORT were first purposively sampled by Hampshire Hospitals NHS Foundation Trust after which a randomiser was applied to ensure the final selection process was free from bias. In February and March 2023, 29 semi-structured interviews (15 EBRT, 14 TARGIT-IORT) were conducted virtually via Zoom. Thematic analysis of verbatim interview transcripts was then carried out by two coders generating 11 themes related to either EBRT or TARGIT-IORT.

A number of procedural grievances were noted among EBRT patients. EBRT was perceived as being disruptive to a range of normal routines (work, home-life, and the burden of travel), and dissatisfying due to discomfort of side effects. TARGIT-IORT was perceived by patients and care partners as being efficient (given while they are asleep during surgery and without additional hospital visits) with minimal if any disruptions to QoL and that it was the safer option. The need for adequate, accessible information provision at the right time to reduce anxieties was noted in both cohorts. These new insights are valuable for healthcare staff and policy makers and could help incorporate the two treatments into routine practice. Further research is now needed to explore TARGIT-IORT in more diverse populations and in the 35 other countries where it is already a well-established treatment option.

## Introduction

Conventional radiotherapy treatment for breast cancer involves patients undergoing external-beam radiotherapy (EBRT) several weeks or months after their lumpectomy (surgery to remove the tumour). EBRT is usually delivered to the whole breast from outside of the body, as the patient lies on a treatment table.

Patients are required to attend treatment sessions, each lasting about 15 minutes, five days-a-week over three-six weeks or, as in some units that follow the new FAST-Forward protocol, administering radiotherapy over five sessions. Additional 5 to 8 days of tumour bed boost is given in about a quarter of cases who are found to have higher-risk disease.

Targeted Intrabeam Intraoperative Radiotherapy (TARGIT-IORT) offers an alternative to women with early breast cancer that is currently being used in a small number of hospitals across England. This approach, first used in 1998, delivers a single dose of radiotherapy directly to the breast tissue surrounding the tumour immediately after the tumour has been removed and the patient is still under the same anaesthetic in the operating theatre. The long-term results of the international randomised TARGIT-A trial (n=2298) in which TARGIT-IORT was compared with EBRT, found TARGIT-IORT to be effective as whole breast radiotherapy, reduced non-breast cancer deaths and improved overall survival in those with grade 1 and grade 2 cancers (Vaidya et al., 2020, Vaidya et al., 2021).

To date, several studies have investigated patients’ experiences with TARGIT-IORT quantitatively (Sosin et al., 2018, Welzel et al., 2013, Corica et al., 2016, Corica et al., 2018, Keshtgar et al., 2013, Andersen et al., 2012). These studies gathered information about patients’ Quality of Life (QoL) during and after treatment via questionnaires and have concluded that patients receiving TARGIT-IORT report high QoL scores (Sosin et al., 2018) and better emotional wellbeing, less pain, fewer breast, and arm symptoms compared to patients receiving EBRT (Welzel et al, 2013). Patient preferences have also been explored in studies based in USA and Australia (Alvarodo et al., 2014, Tang et al., 2021, Corica et al.,2019, Corica et al., 2014). However, qualitative insights can give researchers and practitioners an in-depth of understanding of patient perceptions that can help explain, with confidence, the reasoning behind the difference in quality of life experienced by patients having these treatments.

Much of the literature on patients’ experiences of receiving radiotherapy has focused on EBRT alone where qualitative studies have utilised various methods such as workshops, interviews, and diary entry analysis. Recurring themes include the need for adequate information provision, healthcare professionals’ knowledge of breast lymphoedema (swelling of the arm where lymph nodes were removed), perceived lack of choice, experiences of being naked, and feelings of disempowerment (Probst et al., 2021), psychological burdens of impact (and the resources required to support patients) (Llewellyn et al., 2019), impact of side effects such as skin toxicity on patients’ QoL, life and health after radiotherapy and feeling mystified by radiotherapy and how it works (Schnur et al., 2009, Schnur et al., 2011). While there are other studies investigating breast cancer patient’s lived experiences of receiving the diagnosis, treatment perceptions, experiences of survivorship and symptoms from radiotherapy (Williams and Jeanetta, 2016; Shaverdian et al., 2018; Murchison et al., 2020; Long, 2001; Hickok et al., 2005; Haviland et al., 2016), they do not focus on lived experiences of receiving EBRT specifically.

In addition, the literature review has highlighted that no qualitative comparison of patients’ experiences of TARGIT-IORT and EBRT have been conducted although one qualitative study, exploring overdiagnosis of breast cancer, did briefly describe the experiences of patients having TARGIT-IORT and EBRT (Pickles et al., 2022).

Rich descriptions of authentic experience can help to place the treatment pathway in the context of patients’ *everyday world*, and to truly understand the perceived barriers, benefits, and personal consequences of treatment. Therefore, this study is designed to gather a deep understanding of how patients define the benefits and impacts of each therapeutic regimen and how this qualitatively affected patients and / or care partners’ experiences. As secondary aims, the study will also identify where there have been unnecessary treatment-related impacts on QoL and areas of potential improvements.

## Methodology

### Study design and ethics approval

This study used a qualitative research design with semi-structured interviews as the primary research instrument. Researchers adopted a phenomenological approach which encourages a *bracketing off* of researchers’ own preconceptions and opinions to help mitigate bias and promotes a special importance to *individual* human experience where multiple realities exist (based on participants’ own subjective experiences). At the inception of the study, a patient-led Working Group was established to ensure the research was designed and conducted in a respectful and sensitive manner. Figure 1 outlines the research process. The study received ethics approval from the Health and Social Care Research Ethics Committee B on 1^st^ November 2022. The authors have used the consolidated criteria for reporting qualitative studies (COREQ) to report the study (Tong et al., 2007).

**Figure 1.**
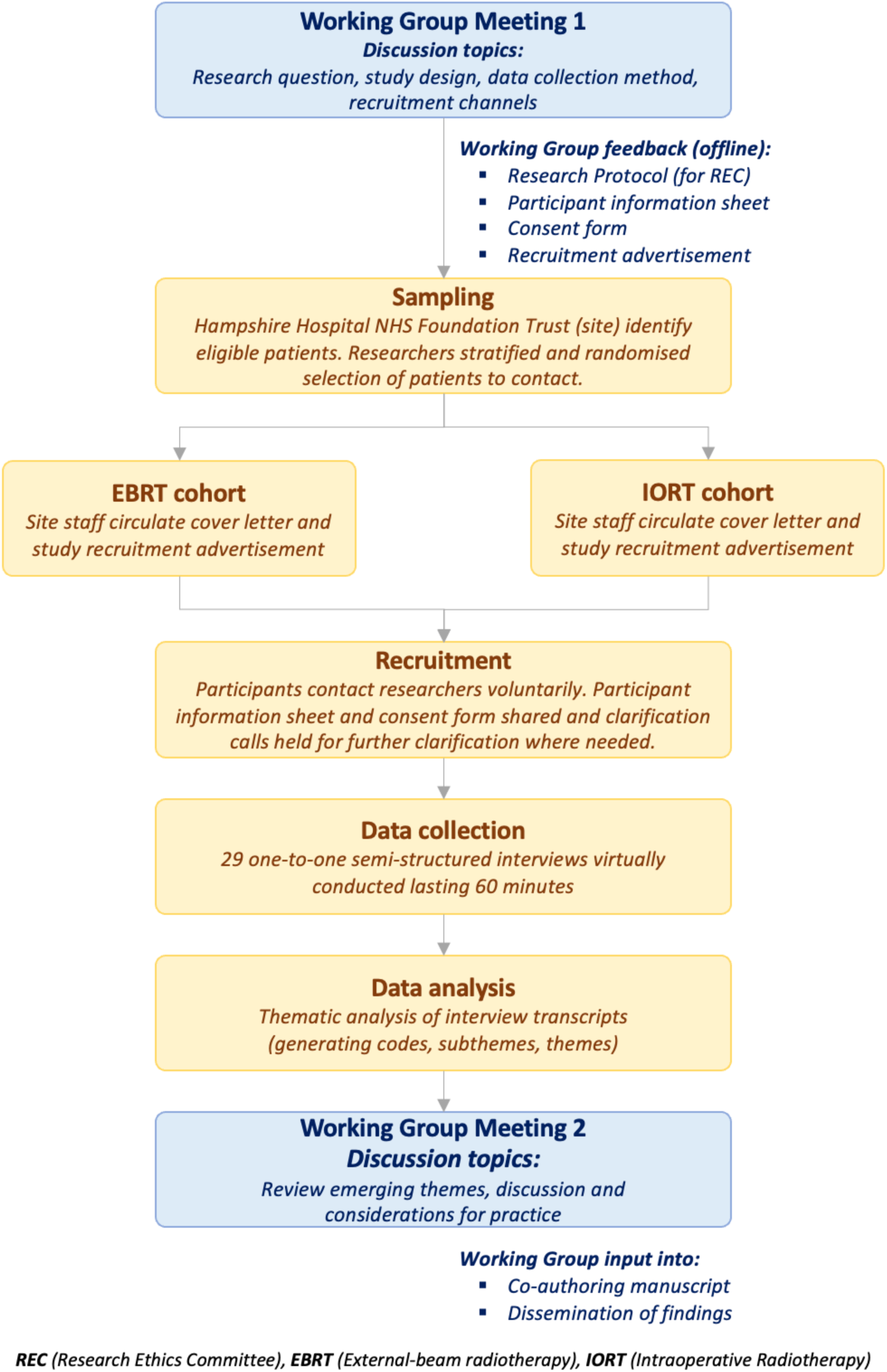
Overview of research process.

### Working Group

Four patient advocates (three patients who had been treated for breast cancer and one care partner) with lived experiences of radiotherapy, were invited to participate in a Working Group with the researchers. An initial meeting was held on 19^th^ August 2022. In this meeting the research design was discussed which included reviewing the study aims, the need for a comparison group, data collection method and participant recruitment channels. The second meeting, on 30^th^ May 2023, focused on the emerging themes from the analysis. Between these meetings, the researchers shared early drafts of the core research material (e.g., participant information sheet and consent form) to obtain members feedback and suggestions for amendments. The Working Group have also co-authored this paper.

### Recruitment of participants and consent

A key outcome of the first Working Group meeting was to ensure the study design had a comparison group. This meant recruiting patients or care partners with lived experience of receiving EBRT to enable a comparison to those that had received TARGIT-IORT. The eligibility criteria are based on criteria used previously in TARGIT- IORT clinical trials (Table 1).

**Table 1.** Eligibility criteria for interview participants.

| Inclusion criteria | Exclusion criteria |
| --- | --- |
| • Female patients aged 50-70<br>• Previously or currently diagnosed with early-stage breast cancer<br>• With experience of receiving EBRT or TARGIT-IORT in England<br>• Care partners (male or female) of female patients<br>• Able to speak English | • Male breast cancer patients<br>• Individuals that are members of potentially vulnerable groups (e.g., prisoners) and / or lack the mental capacity to be able to give informed consent (as defined by the Mental Capacity Act 2005) |

**EBRT** (external-beam radiotherapy), **TARGIT-IORT** (targeted intraoperative radiotherapy)

Participants were first purposively sampled by Hampshire Hospitals NHS Foundation Trust. This Trust recruited patients to the randomised TARGIT-A trial which recruited between 2000 and 2012. Since the National Institute of Health and Care Excellence (NICE) recommendation to offer TARGIT-IORT to suitable patients, they have been offering the procedure to their patients. The breast team compiled a list of all patients that received either TARGIT-IORT or EBRT. Prospective participants were stratified first into rural and urban subgroups and then by age (50-60 and 60-70). A randomiser was then applied to these subgroups to ensure the final selection process is free from bias from clinicians who had treated patients. Cover letters and recruitment advertisements approved by the ethics committee were posted to 58 eligible patients by the clinical team.

Those interested to participate in the study contacted researchers voluntarily. Subsequently the researchers provided further information and shared a participant information sheet and consent form. In total 29 participants were successfully recruited to the study. Participant characteristics can be found in Table 2.

**Table 2.**
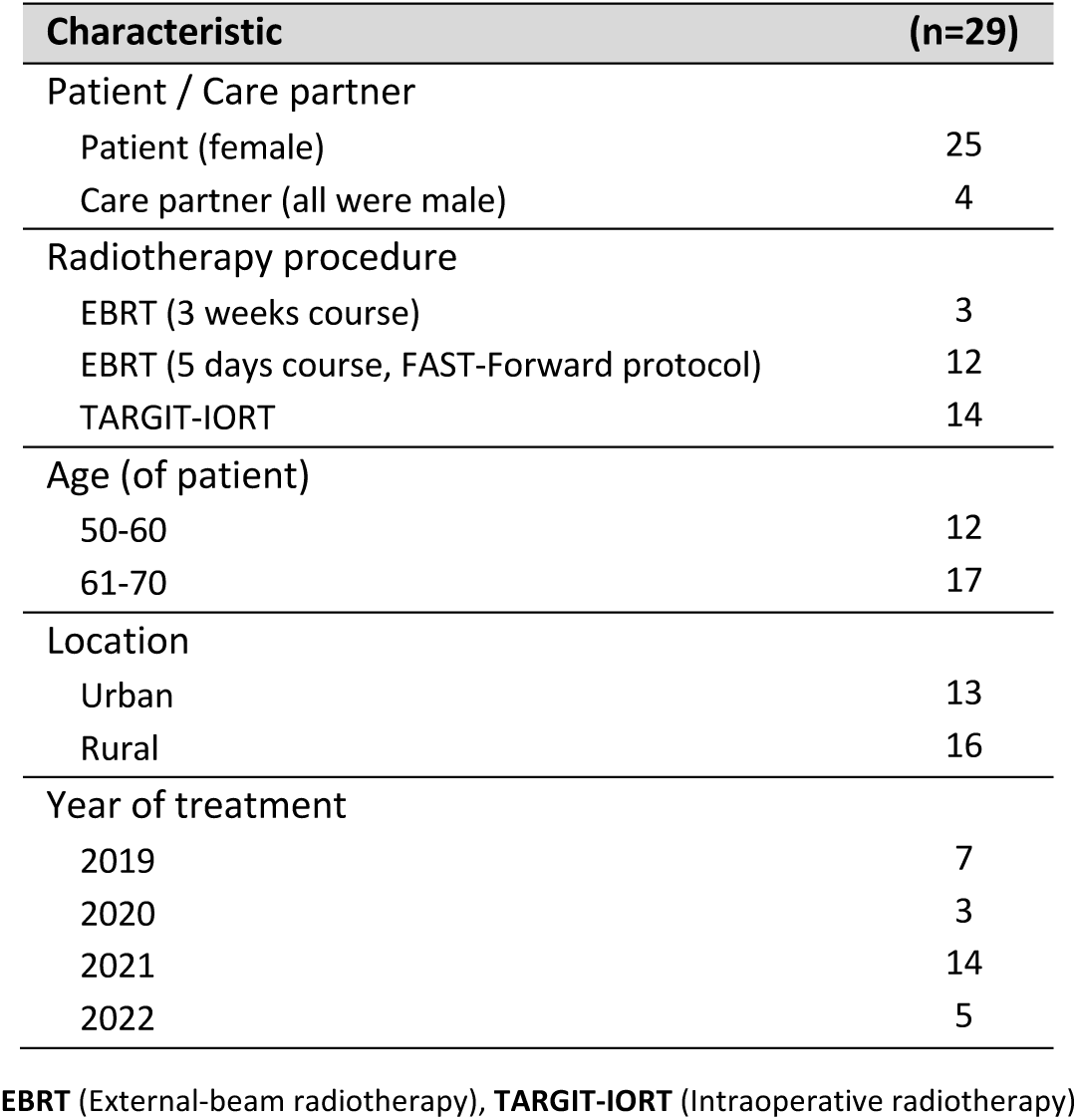
Sample characteristics of interview participants.

**Table 3.** Themes and subthemes arising from the interviews.

| Theme | Subtheme | Study aim(s) |
| --- | --- | --- |
| **External-beam radiotherapy (EBRT)** |  |  |
| *Dissatisfaction with unalterable elements of EBRT procedure* | *Intimidation and discontent* | 1, 2 |
|  | *Dissatisfaction with planning appointment* | 1, 2 |
| *Unanticipated disruptions cause helplessness* | *Delays and extensions cause aggravation* | 1, 2 |
|  | *Covid meant isolation from care partner* | 1, 2 |
| *Straightforward sessions are met with surprise* | *No major complications encountered* | 1 |
|  | *Individual sessions are quick* | 1 |
| *Disruption to normal life routines* | *Patient and care partner work impacted* | 1 |
|  | *Disruption to work has emotional toll* | 1 |
|  | *Travel during treatment is burdensome* | 1, 2 |
|  | *Prolonged disruption to normal routines* | 1 |
| *Experience characterised by discomfort from side effects* | *Side effects and complications are common* | 1 |
|  | *Sessions can be very uncomfortable* | 1, 2 |
| *Specific anxieties about receiving EBRT* | *Stories from friends and family* | 1 |
|  | *Concerns about radiation impact* | 1, 2 |
|  | *EBRT impacted by Covid pandemic* | 1, 2 |
| **Targeted intraoperative radiotherapy (TARGIT-IORT)** |  |  |
| *Perception that TARGIT-IORT is efficient and aggravation-free* | *Quick and one-off feature is appealing* | 1 |
|  | *Ensure minimal mental health impact* | 1 |
|  | *Rare reports of major disruption* | 1 |
|  | *Restrictions and cautions similar to post-op* | 1, 2 |
| *Convenience of performing TARGIT-IORT during surgery is valued* | Less inconvenience / interruption to life | 1 |
|  | Less travel involved | 1 |
|  | Advantages when limiting hospital visits | 1 |
| *Perception TARGIT-IORT is a safe alternative to standard practice* | Acknowledgement of superior clinical profile | 1 |
|  | No awareness / recollection of procedure | 1 |
| *Novel nature of TARGIT-IORT impresses while prompting early caution* | First impressions are positive | 1 |
|  | Concerns as technique is 'novel' | 1, 2 |
| *TARGIT-IORT patients have high information needs* | Lack of prior awareness of TARGIT-IORT | 1, 2 |
|  | Information is critical | 1, 2 |

### Semi-structured interviews

Working Group members agreed semi-structured interviews should be used to gain rich descriptive accounts of experiences with EBRT and TARGIT-IORT. Members felt discussing sensitive and privileged information, namely people’s experiences of receiving the cancer diagnosis and treatment would be more uncomfortable in, for example, a focus group environment. Therefore, two discussion guides (one for each type of radiotherapy) were developed and refined with the help of Working Group members. Interviews, lasting approximately 60 minutes, were conducting between 9^th^ February and 2^nd^ March 2023 virtually through Zoom and either digitally video or audio recorded depending on participants’ wishes. Recordings were transcribed verbatim, with potentially identifying details anonymised, and assigned a unique identifier.

### Analysis

In keeping with a phenomenological approach to analysis, researchers began by writing reflexive accounts. This involved reflecting on their own experiences, preconceptions and assumptions that have the potential to influence interpretations of participants’ accounts. This process helps to create the self-awareness required when attempting to consciously *bracket out* thoughts and opinions that could lead to bias.

Thematic analysis based on Braun and Clarke’s (Braun and Clarke., 2006, Byrne 2022) six step approach was used to analyse the qualitative data and to identify recurring themes related to patient and care partner experience. The process of data familiarisation took place during data collection, transcribing and re-reading the transcriptions. Initial transcripts were individually coded (identifying units of meaning) by two researchers who then reviewed the other’s codes. Through subsequent discussion and reflection, codes were finalised and applied throughout the remaining transcripts. Through an iterative process, codes were further categorised to form subthemes and themes.

## Results

The following section presents findings from the thematic analysis which looked at EBRT and TARGIT-IORT separately and the outputs from discussions of Working Group Meeting 2. Participants’ quotes have been labelled with identifiers (e.g., P1, P2 etc.) not known to anyone other than the researchers (i.e., not the hospital staff, participants themselves or anyone else).

### Themes coming out of interviews with patients who had External-beam radiotherapy (EBRT)

#### Dissatisfaction with unalterable elements of EBRT procedure

The majority of EBRT participants expressed discontent with many of the *standard* elements of the EBRT procedure. Some participants felt intimidated by the size of the room being *“disturbing”* (P22) and the radiotherapy machine being *“scary”* (P19):

> *“…the room that you go into where the machine is, is cold…it could be a bit warmer. Now, some of that could be psychological because you’re in a big white room with a big, huge machine…” (P3)*

> *“…the 2 nurses go into another side room, so, you feel so alone, and you know, and this machine sort of moving around you. It’s, it is quite scary to deal with. (P19)*

Four participants also described the challenge associated with needing to hold one’s breath during sessions. This is done with the hope that the heart may receive less radiation by pushing the chest wall and the breast away from it. Participants described it as saying, *“that was the worst bit”* (P12), *“it’s going to be difficult”* (P15), “*I don’t want to be zapped on my heart” (*P21) and another felt it was *“really claustrophobic”* (P19) or causing *“panic”* (P19). The planning appointment required for EBRT was met with similar dissatisfaction. While there is a clear appreciation for healthcare staff and their workload, participants were unhappy with the dehumanising nature of these appointments:

> *“You become another face… you do feel like a slab of meat while they’re trying to get you in the right position and it’s not a pleasant experience. (P19)*

These experiences resonated with Working Group members’ recollections: *“silent”* and *“cold, dark room”* and finding it difficult, a *“physical challenge”* to maintain position after surgery. Another member felt that while healthcare staff were pleasant, the experience of receiving radiotherapy itself is *“quite traumatic”* and emotional, *“I remember lying there and tears came from nowhere…”*.

As with study participants, Working Group members acknowledged that while healthcare staff themselves are not at fault, the *“system”* causes the dehumanising elements described by participants referring to poor staffing levels and a high-pressure work environment within healthcare.

In contrast, one participant positively describes the EBRT sessions, *“there’s music on, and I didn’t find any cause to worry at all”* (P15). The relaxing effect of music was echoed by a Working Group member who also recalled how music helped in which she felt was *“brilliant”* and stated, *“it helped my head”*.

Finally, a third of participants expressed a strong objection to being tattooed which was required to ensure radiation is delivered to the right location.

> *“What really did wind me up actually, I had to have the 3 dots tattooed on me and I didn’t want tattoos.” (P3)*

Participants were frustrated by the fact that it is permanent, the colour used, and two participants felt it affected their confidence in wearing certain clothes, *“I can still see that now, if I wear a swimsuit or something” (19).* One Working Group member felt that patients are often uninformed about radiotherapy and that patients’ preferences are not listened to. She concluded that this was a good example of an area that required adequate information sharing at the right time.

#### Unanticipated disruptions cause helplessness

A third of EBRT participants experienced either delays on the day of the EBRT session or extensions to their course owing to either machine failure or staff absences. The impacts on patients and care partners include stress, aggravation, and disappointment with knock-on consequences proving to be burdensome:

> *“…the machine broke down…but I couldn’t find [the new hospital], and I got really tired and upset. I was trying to find where I was supposed to go, and nobody seemed to know, and I just managed to grab the team before they went home. I was like, ‘Give me my last radiotherapy now!’ It was down in some basement I mean. Location S is a maze. So, that was a bit stressy.” (P1)*

Patients that received their EBRT during the Covid pandemic were unable to have their care partners (in most cases husbands) with them at key treatment stages. This isolation caused additional anxieties in the EBRT cohort as one care partner stated, *“it’s disappointing and it would have been nicer for me to be able to support her more…”* (P23). Similarly, a patient participant states:

> *“…it’s anxiety level of just having that, that security blanket of having somebody there with you.” (P19)*

The Working Group discussed isolation and the emotional impact during EBRT sessions. Since EBRT continues for days and indeed weeks in some cases (five-six weeks for all members), patients truly feel alone during this phase. They recalled overwhelming feelings of sadness during the sessions with thoughts such as, *“how did I get here”*. One member expressed empathy with study participants who would have had to endure further isolation during the Covid period.

#### Straightforward sessions were met with surprise but travelling for EBRT was still a concern

Four out of the 15 participants interviewed from the EBRT cohort, stated that no side effects or other complications were experienced with one participant saying she felt like *“one of the lucky ones”* (P20). In the absence of side effects such pain, burning or tiredness, patients experience a sense of surprise and relief after having received prior warnings from either clinical staff or hearing stories from friends and family.

> *“…they said to put a couple of tubes of aloe vera, and keep in the fridge, and put it on religiously. But I was fine. I was fine. I literally had no burning. No rash, no nothing. I’d heard about friends having burns, but I just didn’t, I didn’t.” (P12)*

Similarly, three participants were appreciative of the fact that *individual* sessions of EBRT were in fact *“very straightforward”* (P1) and quick:

> *“I did find the time, actually, went quite quick. It wasn’t very long; it might have been an hour. Yeah, it wasn’t too long.” (P1)*

Although these four participants did not experience radiation related side effects, it should be noted that two of the four participants did, express frustrations about the burden of travelling during EBRT sessions and that the *overall* radiotherapy course was *“time-consuming”* (P2).

#### Disruption to normal life routines

A few participants were either employed or self-employed and described how EBRT impacted on their own work performance (e.g., tiredness, weakness in arm) with one person concluding, *“I’m an office worker but if I’d be doing a manual job, I think it might have impacted more.”* (P3). There is also the realisation that patients would likely need to adjust work sessions to accommodate for the EBRT sessions and the related side effects: *“I’ve worked out a part time basis to get back into work.”* (P19).

Participants shared concerns about the impact on work colleagues. One participant states: *“I’m the only person that does my job. So, I was acutely aware that when I’m not at work other people are picking up my job”* (P19). Similarly:

> *“…we were a short-staffed team, I was aware that when I wasn’t there, it was putting work onto other people, and I felt I should have been there….” (P3)*

The EBRT cohort also revealed the emotional toll of work-related worries and impacts, and the work-guilt associated with the impact on colleagues. Similarly, potential impact on the care partner’s work is also, clearly, a significant consideration for patients:

> *“The first night after my first session I was in so much pain, I mean I didn’t sleep a wink the first night. It was absolute agony…it was my self-confidence, and everything was destroyed…and I didn’t dare, I didn’t want to wake my husband up. He had to go to work early, so then he could take me to radiotherapy in the afternoon.” (P19)*

These impacts on employment were corroborated during Working Group discussions. One member, commenting in particular on self-employed people, described the impact on financial standing and home life as *“catastrophic”*.

In contrast, those that had flexible hours of working were less affected, as one patient participant describes her care partner:

> *“luckily he was doing a job where could sort of pick and choose what he did and his hours, so it was all right.” (P3).*

The repetitive nature of the EBRT sessions, the travel involved, and the side effects experienced all also impacted on participants’ *normal* daily routines. For instance, home activities such as shopping, gardening and caring responsibilities were impacted:

> *“I think we might have cut down [caring for elderly parents] to once a week instead of two or three times…” (P3)*

One participant mentioned a close family member taking a week off from work to support looking after her, her husband, family pets and *“doing a few household chores like pushing the Hoover around”* (P22).

Severe pain brought on by EBRT was described as *“agony”* (P19) both during sessions and after sessions (P7). Ongoing pain months and years after the radiotherapy means patients need to settle into a new norm, now constantly having to be aware of their own *“limitations”* (P6):

> *“I’m still aching like mad from [radiotherapy], that’s two years later and I’m still achy and in pain.” (P6)*

For Working Group members, these experiences were familiar. One member clarified that although there were no specific side effects from the radiotherapy the disruption to life was *“hugely problematic”*.

Many of patients who received the FAST-Forward protocol included sessions either side of a weekend resulting in a total of seven days to complete the course. No participants complained specifically about the weekend being involved in this way however many described dedicated activities for the weekend for example, food shopping, family commitments and short trips away which would undoubtedly be affected while in active treatment.

Conversations with participants revealed personalities play a role in patients’ cancer journey. One participant felt the emotional impact of radiotherapy treatment (and the subsequent withdrawal of routine contact with healthcare staff) more than all the others. The anxieties associated with this perceived void and the mental health impact itself is disruptive to reengaging with normal life and routines:

> *“…you’ve got people checking on you as in consultants or the breast care specialist nurses, your GP, the radiographer, people ringing you, checking on you, you’re seeing them all the time. After the radiotherapy, I’m suddenly though on my own now. I didn’t realise it was a real security blanket…it was so reassuring to seeing the consultant and seeing this nurses every day. That was the bit where I took a bit of a dive…I had two or three days where I couldn’t stop crying, I thought, ‘Oh I’m on my own now’…” (P12)*

There were also those who demonstrated a *pragmatic mindset* viewing any discomfort and impactful delays associated with EBRT sessions as realities to, in effect, take in their stride, “*any inconvenience, you just get on with it.” (P20)*

Travel is clearly an uncomfortable reality and demanding aspect of receiving EBRT. Many participants complained about the repeated journeys required for EBRT sessions. The burden has been described as *“dreadful”* (P7) with people feeling *“exhausted”* particularly where the effects of radiotherapy (tiredness and pain) are felt. The burden of travel also manifests as experiencing a longer day overall as well as sheer cost of public transport used to make the trips independently:

> *“It’s a pain having to go to Location S, there’s a hospital there where the bus doesn’t even go. So, from Location O, I have to get a bus from here to Location H, then wait half an hour, then from Location H to Location W, bus station, then a taxi to the hospital. That all costs 30 quid and is time-consuming, and when you’re sore, it’s not ideal.” (P2)*

In the majority of cases, there is a reliance on others (the husband but on occasion friends or children) to drive participants to the EBRT sessions. While there are no direct statements indicating a burden to care partners, it is important to note that in these cases both the patient *and* the care partner endure the repeated journeys:

> *“My husband drove…I’m not a good driver…I certainly can’t park so it was good that he went with me because in case, if something went wrong or something because he’s like my rock he is.” (P3)*

The location of the EBRT sessions is critical to the quality of patient experience and when participants were presented with two hospitals to choose from it was clearly valued:

> *“…it’s the same surgeons, all the same team, who were in Hospital W or B. I opted for Hospital B, it’s nearer to me and I could get to Hospital B easily, whereas Hospital W was an ordeal for me.” (P21)*

Two participants that were retired and for who the location of the hospital was particularly close indicated that travel was *not* a burden and considered themselves *“very lucky”* (P22), though of note, one of these participants experienced no EBRT side effects which may have contributed to relatively positive experience.

In contrast to many of the experiences described above, one Working Group member shared that she was relieved to have regained her independence since she was able to travel to her EBRT sessions herself.

However, the Working Group notes that all their experiences included a difficult period of receiving chemotherapy first where radiotherapy was viewed as the *“lesser of evils”*.

#### Experience characterised by discomfort from side effects

A wide range of side effects, clearly attributed to EBRT, were reported by the vast majority of participants and this characterises an important part of the EBRT experience. These included varying intensities of: tiredness; burning (from warm sensation to blistered and sore); skin-related conditions (dryness, itchiness, rash); pain; and breast size and density changes. The most common complaint was tiredness:

> *“…there were 3 or 4 days the following week when I just had to go to bed, or just have to, you know, lie on the sofa in the afternoon, or you know I was just really bombed out, and I’m not someone who goes to bed and afternoon normally, I’m always busy, and but I had to. I was knackered.” (P12)*

EBRT sessions can be very uncomfortable. Severe pain, described as *“agony”* (P7, P19) was experienced by two participants. One individual, unable to withstand the pain and the tiredness experienced, made an independent decision to stop attending the sessions for a few days:

> *“I was very tired…I think I might have missed a few days, because I couldn’t make it in between…I thought I’ve done so much now, I’m not going to go anymore because it was really, really hurting…I just wanted it to end and go away? And not think about it anymore basically.” (P7)*

Patients experienced some side effects such pain in the breast or weakness in the arm for a prolonged period – months and years after the EBRT sessions. There were also reports of new symptoms requiring follow up that were attributed to EBRT. One participant (P8), who felt uninformed about the *“long-term, lasting and late effects of radiotherapy”* had developed a new pain under her ribs – she reflected:

> *“I would not have had radiotherapy and I would not be beating myself up about having had it now had I been given the full information about the long-term effects. You know, it’s life-shortening, radiotherapy is life shortening in itself like chemotherapy.” (P8)*

During discussions at the Working Group Meeting 2, members were not surprised by the insights captured from study participants and felt strongly that there were *“no positives”* from EBRT (particularly when compared to TARGIT-IORT). One member reflecting on their experience with EBRT stated they came away thinking “*almost anything is better than this.” (WG member)*

#### Specific anxieties about receiving EBRT

Discussions with the EBRT cohort revealed three main anxieties associated with receiving radiotherapy. Firstly, while there is evidence that more information early on (particularly from consultants) helps to reduce worry, stories of radiotherapy experience from friends and family members can raise concerns and anxiety levels:

> *“I only knew what I knew as a lay person, you know, various friends have had radiotherapy, unfortunately, people know a lot of people who have all these sort of things… I’d heard about friends having burns…”” (P12)*

> *“…and actually, talking to another friend, she said she would do chemotherapy any day over radiotherapy because of how the radiotherapy, the pushing around and making you feel like a piece of meat, how it how it made her feel.” (P19)*

The second concern was the potential for radiation to cause harm: (a) so soon after surgery; and (b) to healthy tissue and organs: *“You’ve got to heal a bit; you can’t go straight into radiotherapy because obviously you’re as raw as hell*” (P6). Two participants in particular experienced discomfort in their arm during their EBRT sessions – one participant had a number of lymph nodes removed from the armpit and one participant developed a seroma which is an abnormal accumulation of fluid following surgery. A Working Group member shared a similar experience where the development of seroma delayed the start of her radiotherapy course.

Another participant who was particularly concerned about unnecessary radiation exposure requested that only half the breast was irradiated because she wanted *“the absolute minimum”* (P8). Similarly, another care partner described his wife’s concerns:

> *“One thing is that my wife was worried about was the radiotherapy because obviously there is this thing with radiotherapy, particularly on the breast, of potential damage to the lungs and she was very concerned about that.” (P23)*

Thirdly, since many of the participants received their EBRT during the Covid pandemic, a few expressed worries about potential delays either due to staff shortages, protocol-driven cancellations (i.e., limiting patient numbers) or themselves contracting Covid (since multiple visits are required with EBRT) and thereby being unable to attend hospital:

> *“But covid was going on and I remember being so scared that my appointments would be cancelled.” (P12)*

### Intraoperative radiotherapy (TARGIT-IORT)

#### Perception that TARGIT-IORT is efficient and aggravation-free

Of the 14 TARGIT-IORT participants interviewed, 11 indicated the *one-off* feature of the procedure was appealing. There are many references to how quickly the procedure was completed *“it’s lovely to get it all done and finished with on the day”* (P26). Similarly:

> *“…having it done and dusted, and then then waving goodbye at the hospital gates, it was like why, why would I say, ‘No thank you’?” (P16)*

As a consequence of this efficiency, there is relief that the procedure permitted radiotherapy to be administered without any Covid-related delays, or exposure to the COVID-19 virus during travel or hospital during the multiple visits for EBRT, or complications for which three participants had expressed initial concerns:

> *“I was just delighted that it was dealt with really, really quickly, because back at that time the news was full of things where, you know, because of Covid, you know, everything has been delayed, and people not getting cancer treatment and that was one of my, I remember having that conversation with the consultant and said, ‘Look are we going to be delayed’.” (P25)*

There is a similar relief detected in participants discussing the potential positive impact TARGIT-IORT can have on patients’ mental health as one care partner states:

> *“…the alternative would have been [EBRT]… her symptoms of depression are she gets very, very tired… so intuitively our reaction to [TARGIT-IORT] was… actually quite a good idea.” (P24)*

Going through a cancer diagnosis and receiving treatment was clearly an emotional time. One participant was impressed with TARGIT-IORT precisely because the efficient delivery of radiotherapy facilitates her moving on quickly:

> *“…the beauty of intraoperative radiotherapy is that I could say ‘OK, been there, done that, move on.” (P9)*

#### Convenience of performing TARGIT-IORT during surgery is valued

Most participants from the TARGIT-IORT cohort shared why they preferred to receive radiotherapy at the same time as the surgery. There is a recognition of the convenience that TARGIT-IORT brings as a result of not having to attend hospital on multiple occasions e.g., less travel and car parking and supporting independence (particularly for retired individuals):

> *“It’s my choice to have [TARGIT-IORT] because I thought that it was a better option for me particularly because I live on my own and it would allow me to be more independent.” (P18)*

While the majority of participants were retired, those that did have young children felt TARGIT-IORT supports their caring responsibilities: *“I’ve got a [child] and I’ve got to look after him… This is a better way to go…”* (P14). Additionally, one retired participant who valued the independence TARGIT-IORT facilitated concluded it would also suit younger, busy, women well:

> *“…particularly for younger women this would be an extremely good thing, if they’re working, it allows them to get back to work without that constant interruption and if they’ve got a young family.” (P18)*

Many participants were able to draw from stories and experiences they had heard from friends and families. The apparent inconvenience and impact of daily radiotherapy doses discouraged patients from EBRT when TARGIT-IORT was presented as an option. One participant whose father received daily doses for prostate cancer felt she would *“rather get it all over in one go”* (P10). Similarly:

> *“[TARGIT-IORT] was perfect, because it just meant I didn’t have to queue up in the car park with the other poor people having radiotherapy, and I did have friends who had serious cancers who were having radiotherapy at the time, and it was just miserable.” (P9)*

There is also a perception that with TARGIT-IORT recovery times are likely to be faster since it would be signify the end of their cancer treatment: *“I’m going to get [TARGIT-IORT] and it’s done”* (P16) and *“I can just then get on and recover”* (P4). Another participant summarises her main reasons for opting to receive TARGIT-IORT:

> *“So, there were probably 3 reasons I went for [TARGIT-IORT]. You know, Covid, convenience, and the fact that I thought, you know, ultimately, I’d probably recover quicker.” (P9)*

Only one participant from the TARGIT-IORT cohort, a care partner, described significant logistical impact due to his wife’s cancer treatment in general:

> *“…created quite a challenge really for me, I mean, I was never going to moan about it, I wasn’t the one who just had cancer surgery! But you know, it meant the days suddenly got very challenging…” (P25)*

#### Perception TARGIT-IORT is a safer alternative to standard practice

Five participants felt that they did not experience any complications as a result of TARGIT-IORT and were able to resume their normal activities quickly. While there are a few cases of soreness and itchiness that participants *specifically* attributed to TARGIT-IORT, most participants did not report the range of side effects seen in the EBRT cohort. As a result, participants gave their endorsements for TARGIT-IORT, respectively:

> *“I moved around, I got up, got changed, got dressed. It was surprising actually, this is why I’ve decided to do this, if this is what it gives you then everyone should have it. You know you don’t need to feel debilitated, and you can carry on with your life. I’ve got a [child], and I’ve got to look after him. So, if you can, why not. This is a better way to go if the prognosis allows it.” (P14)*

> *“There were no, no after-effects, no problems. It all healed up very well, because it was quite a small incision anyway and very, very successful.” (P28)*

The majority of participants felt the procedure prevented healthy tissue and organs from being unnecessarily exposed to radiation because *“the radiotherapy is directed immediately where the lump [is]”* (P17).

> *“I confess I heard that and thought ‘God, that’s a bloody good idea, why don’t they do that more often?’. Because obviously if you don’t have to beam through loads of flesh and muscle to get at what you’re aiming for then that’s got to be better to be honest.” (P24)*

A few participants described side effects (soreness, tiredness), precautions (new bra needed, seatbelt cushion) and restrictions (no pressure, sport, lifting) however they were unable to clearly attribute whether these were related to the surgery or the TARGIT-IORT procedure since both occur at the same time.

> *“…yeah, my arm was a little bit sore…I’m sure it must have been the radiotherapy or the operation, I don’t know.” (P29)*

> *“…a special seat belt cushion that protects your breast from the seat belt and I had one another cushion under my breast supporting it…” (P11)*

#### Novel nature of TARGIT-IORT impresses while prompting early caution

Although it has been in use for the last 25 years since the first case was done in 1998 TARGIT-IORT is seen as novel and innovative with advantages acknowledged over EBRT. The decision to proceed with TARGIT-IORT is widely considered *“easy”* (P28) or *“intuitive”* (P24) or a *“no brainer”* (P4):

> *“…well, you’re in there, so you might as well get on and do it and that would surely save the need for me having to come back, I can then just get on and recover basically…it was a no brainer for me, an absolute no brainer.” (P4)*

However, a few participants described their initial concerns since TARGIT-IORT was introduced by the consultants as a clinical trial and was largely unheard or *“unknown”* (P9). Care partners, often husbands and sometimes participants’ children wanted to carry out their own research to help making an informed decision about TARGIT-IORT. One participant had already felt she was convinced by the consultant’s explanation and the advantages over EBRT however her daughter, who worked in healthcare, stated *“…‘hold on a minute, we need to look at the statistics and the recovery times, side effects’…”* (P10). Similarly, another participant’s husband wanted to an opportunity to ask the consultant more questions to help feel more reassured:

> *“…but [care partner] just wanted to have the conversation around the intraoperative radiotherapy because it was an unknown really.” (P9)*

It should be noted that many of the participants were either themselves or their close family (e.g., husband) highly educated, often with a science-based background, and were able to explore clinical study papers and statistics: *“I’ve got a little statistical training…so I looked at the stats and what the mean variation was…what the levels of certitude at either end of the scale were…”* (P24).

#### TARGIT-IORT patients have high information needs

As mentioned above, due to the relative novelty of TARGIT-IORT and in the absence of experiences of TARGIT- IORT among participants’ friends and family, reliable information from trustworthy sources is critical. The majority of participants (in both EBRT and TARGIT-IORT cohorts) displayed high levels of trust in their consultant. Receiving adequate information from them about TARGIT-IORT, particularly due to its initial availability via a clinical trial, was appreciated:

> *“I think what was good was the way that it was explained in the first place and what the pros and cons were, or if in fact, there weren’t any cons really at all…So, you know, we were told that the treatment, doing it during the operation, is just as effective but it would mean that you would have no subsequent radiotherapy and, you know, of course I’m young and foolish, I assume that to be true, we trust the doctor…” (P25)*

> *“[The consultant] said’ ‘This is this, that is that…pluses and minus’…gone through pros and cons and I had made up my mind that that was a good way to go.” (P14)*

Working Group members could relate closely to this subtheme of *trust.* They explained that the retrospective perception of TARGIT-IORT was always likely to be a *“no brainer”* however for a patient going through the highly emotionally-charged process of receiving their diagnosis and treatment, at a time when they are already overwhelmed with new information, the relationship with the doctor is important:

> *“…if it’s being offered to you, it’s important how it’s being offered to you. We put out trust in, so much, our doctors.” (WG member)*

Two participants described receiving explanations from radiation oncologists during their pre-surgery appointment, however, these discussions were not influential in helping to decide which type of radiotherapy they would receive. A few participants were wary of using the Internet to search for information related to their treatment options: *“I’m very cautious of what information I take in from Google”* (P4). However, the majority did conduct their own Internet searches to bolster their understanding of TARGIT-IORT:

> *“I then went away and looked the bugger up, and then you could learn for yourself a little bit, reading between the technical stuff, what it’s all about, the success rate is there or there about the same, it’s not wonderful but for me, it was a no brainer. (P16)*

The provision of information was discussed on a number of occasions by Working Group members. Simple and clear language is particularly important at a time when patients are already in a vulnerable, stressed, and emotional state:

> *“…you are so blindsided…the normal way you operate doesn’t necessarily apply.” (WG member)*

Working Group members pointed to the need for information sources to be created adequately in the first place, for example, being written by patients / care partners who possess the lived experiences and so are able to elaborate on the areas that matter.

> *“…there should never be a need for a patient to go home and want to Google, you should go home with the information in hand or go home with reputable evidence-based sources of information.” (WG member)*

## Discussion

The primary finding of this study is that the subjective experiences of patients and care partners receiving EBRT or TARGIT-IORT differ significantly. Strong recurring themes of appreciation and recognition of innovation, convenience, absence of side effects and lack of disruption to life have emerged from the TARGIT- IORT cohort while in the EBRT cohort, we have largely heard about discomfort and disruption to life. Notably, EBRT patients experienced disruption to their life despite the majority (12/15) received their EBRT with the shorter, compressed Fast-Forward regimen. We can expect that this disruption would be much greater using the well-documented and less toxic 3-week regimen. These themes – centring around: (a) treatment procedure itself; (b) impact on QoL; and (c) information needs – was presented to and were validated by a patient-led Working Group.

Patients and care partners involved in this study described numerous challenges, concerns, and dissatisfaction with elements of the EBRT procedure while processing a difficult and emotional diagnosis. These findings are consistent with the existing literature on EBRT experience. Probst et al. (2021) also identified procedural grievances, for instance, patients described the radiotherapy sessions as *“dehumanising”*, *“emotionally draining”* and complained about the tattoos being a permanent reminder of the cancer. Previous studies exploring patient-perceived barriers to radiotherapy include patients’ fear surrounding radiation toxicity which can result in non-compliance and insufficient treatment (Guidolin et al., 2017; Sosin et. al, 2018). In fact, research has identified that fears and anxiety regarding the EBRT experience can influence a patient’s decision to opt for a mastectomy over EBRT, despite the latter having equivalent if not non-inferior survival rates (Shaverdian et al., 2018). Several studies have demonstrated that as the distance from radiotherapy centre increases, the rate of mastectomy also increases (Goyal et al., 2014, Athas et al., 2000, Nattinger et al., 2001, Celaya et al., 2006). Indeed, this was the primary patient-centric reason that the TARGIT-IORT procedure was originally conceived (Vaidya et al 1996, Vaidya et al 2001, Vaidya et al., 2010).

Our study demonstrates the need for improvements in the way EBRT is delivered and has implications for practice that extends to cases where patients are not eligible for TARGIT-IORT. In stark contrast, those receiving TARGIT-IORT have no awareness or recollection of the procedure since radiation is administered during surgery. Patients and care partners found this feature particularly appealing and contributed to their decision to opt for TARGIT-IORT. Indeed, TARGIT-IORT has been widely adopted elsewhere and treated 45,000 patients across 38 countries (Vaidya et al., 2022).

It is evident that QoL-related benefits and impacts are a central component of radiotherapy lived experiences. Compared to TARGIT-IORT, EBRT has a prolonged impact on patients and perhaps a compounded impact on QoL where patients live alone (lack emotional or practical support), don’t drive (reliance on others or public transport with additional costs and travel time) or have caring responsibilities (partners, parents, children, pets). Travel and mobility issues have been recognised as barriers already (Guidolin et al., 2017) as has the inconvenience of a prolonged treatment plan which can affect those living in remote areas even more (Sosin et al., 2018). Our findings demonstrate the advantages TARGIT-IORT offers to those that are eligible. All participants in our study acknowledged the efficiency of the procedure with many drawn to the option (over EBRT) because it was considered *“straightforward”* and *“over-and-done”* during surgery. The benefits of TARGIT-IORT to patients in terms of cost, travel time and distance have been demonstrated, in principle, elsewhere (Bargallo-Rocha et al., 2017). Furthermore, the environmental and social impact of the substantially more travel required for EBRT, and huge reduction in carbon-footprint from cancer treatment by use of TARGIT-IORT has also been well documented (Coombs et al., 2016).

In our study, inconveniences and logistical complications were exacerbated by EBRT side effects which were recognised as a key characteristic of the EBRT patient experience and have implications on QoL. Stanton et al. (2001) investigated factors affecting QoL during and after radiotherapy and found that functional impacts of treatment, particularly breast specific pain (on e.g., mobility) are important correlates of QoL. In addition, Schnur et al. (2009) showed that key patient concerns include the timing of side effects and the impact of side effects on self-esteem affecting patients’ perception of being attractive, good workers, patients, and parents. Another study supports these findings and also shares one case of such extreme physical discomfort (pain, burning etc.) that the patient admitted she had considered ending treatment and another saying she would never choose to have radiotherapy again due to the burning sensation (Schnur et al., 2011). Our study has captured similar cases. This study also underscores the emotional toll, anxieties, and stresses that disruptions to life (e.g., work-related) cause and have been heard at a NICE Committee meeting (NICE., 2018). There were fewer reports of side effects *directly* attributed to TARGIT-IORT in our findings. This is consistent with a study comparing TARGIT-IORT with EBRT (quantitatively) in which patients receiving TARGIT-IORT also reported less pain, fewer breast, and arm symptoms, and better every-day functioning when compared to patients receiving EBRT (Welzel., 2013). However, our study detects apparent patient *obscurity* between surgery-linked or TARGIT-IORT-linked side effects – this clearly needs to be addressed through appropriate education and adequate information provision.

Results from our study cohorts point to the need for improvements in communication and information provision. The role of high-quality communication by healthcare staff and access to emotional support services, particularly when radiotherapy treatment ends has been highlighted already (Llewellyn et al., 2019). Previous research has also identified that patients can often feel mystified by radiotherapy (EBRT), how it works and will have anxieties about life and health after radiotherapy (Schnur et., 2009) or feel disempowered and lacking the ability to make an informed choice (Probst et al., 2021). In our study, Working Group members emphasised the importance of *trust* in connection with information provision particularly during an emotional cancer diagnosis. Members felt a number of study findings could be addressed adequately by effectively communicating the right information at the right time. Examples include: letting participants know clearly that tattoos will be permanent; what the immediate and long-term side effects of both radiotherapy types are; understanding the side effects of surgery thereby avoiding confusion with TARGIT-IORT; ensuring TARGIT-IORT explanations are always supplemented with lay language overviews of the efficacy and safety profile compared to EBRT. One study showed that more than 90% of patients felt that if they were more informed about radiotherapy, they would be less scared about it (Shaverdian et al., 2018). The Working Group advocated for any shared information, such as leaflets, to be written *by patients* i.e., those that have experiences of receiving radiotherapy and therefore have an awareness of where there are likely to be challenges in understanding treatments and their impacts clearly. Similarly, considerations also ought to be given to ensuring people with learning disabilities and communication difficulties are able to make an informed choice by developing *accessible* information. To satisfy ‘valid consent’, doctors in the UK are now obliged to follow the new GMC guidelines underlining the essential nature of adequate patient information (GMC, 2020) about all proven treatment options, even if they are not available at their own centre. In the UK this powerful principle is now fully enshrined in law (Montgomery v Lanarkshire Health Board, 2015). The substantially better patient perception and experience documented in this study needs to be included during consultations with patients when discussing treatment options before they have their surgery for breast cancer.

### Strengths and Limitations

A major strength of this study design is the inclusion of patients who received TARGIT-IORT as well as those who received EBRT, allowing assessment of patient’s perceptions in each of these two types of treatments. Another strength of this study is that measures were put in place to mitigate researcher bias. Researchers each wrote a reflexive account to help maintain an objective standpoint and interview transcripts were independently coded by two researchers with rigorous discussion to help generate an unbiased coding frame. Where possible multiple perspectives have been sought – firstly, through capturing the views and experiences of care partners and secondly those receiving current standard of care (EBRT) to enable a comparison.

Furthermore, the study population included a good mix of age and urban-rural areas. Finally, our involvement of a patient-led Working Group has supported efforts to design a study that is conducted sensitively and robustly and helped to validate our findings.

The limitations are that the study population was from just one location in England and, most participants are white and belonged to higher socioeconomic groups. Further research is needed to explore the lived experiences of radiotherapy in early breast cancer in more diverse populations in England and indeed other countries where TARGIT-IORT is an established treatment option.

Many of our participants received their cancer diagnosis and treatment during the COVID-19 pandemic. While this has helped to highlight issues such as patient isolation in EBRT recipients and its absence in TARGIT-IORT recipients, it could also be seen to add obscurity to the some of the findings.

## Conclusion

This qualitative study, co-led by patients, uncovered detailed lived experiences of patients treated for early breast cancer (and their care partners) receiving either EBRT or TARGIT-IORT. The research demonstrated a patient-perceived superiority of TARGIT-IORT over EBRT – it is considered more efficient with less disruption to life routines. The paper also illustrates the importance of provision of accessible information about all radiotherapy treatment options from trusted sources, at the right time (before breast cancer surgery), to reduce initial anxieties and help patients make informed choices. These new insights need to be taken together with the established quantitative survival and QoL benefits of TARGIT-IORT over EBRT. We believe that these deep insights into the patient’s perspective will substantially improve our understanding of the lived experiences of patients with breast cancer and will help clinicians, patients, and policy makers to comprehensively consider how access to better treatments in the NHS can improve patients’ lives. Further research is now needed to explore TARGIT-IORT in more diverse populations and in the 35 other countries where it is already a well-established treatment option.

## Data Availability

All data produced in the present study are available upon reasonable request to the corresponding author.

## Author contributions

SB, NS, CH, MEO, SF, DW and RW were responsible for the study concept and design. SB, NS and JR collected the data. SB and NS analysed and interpreted the data. SB and CH wrote the first draft. All authors were involved in reviewing draft iterations. MEO, SF, DW and RW approved the report. The authors take full responsibility for the report.

## Acknowledgments

The trial was initiated by MediPaCe, a patient engagement and patient research company working with the industry. The manufacturers of the Intrabeam device (Carl Zeiss Medtech AG) did not have any part in concept, design, or management of the trial, or in data analysis, data interpretation, or writing of the report. We thank Prof Jayant S Vaidya for reviewing the draft manuscript and contributing to its intellectual, contextual and factual content. The study was sponsored by MediPaCe. Funding was provided by Carl Zeiss Medtech AG.

## References

1. Alvarado, M., Conolly, J., Park, C., Sakata, Th., Mohan, A., Harrison, B., Hayes, M., Esserman, L., Ozanne, E. (2013), ‘Patient preferences regarding intraoperative versus external beam radiotherapy following breast-conserving surgery’, Breast Cancer Res Treat, 143(1). p 135–40. Available at: https://pubmed.ncbi.nlm.nih.gov/24292868/

2. Andersen, K., Gärtner, R., Kroman, N., Flyger, H., Kehlet, H. (2012), ‘Persistent pain after targeted intraoperative radiotherapy (TARGIT) or external breast radiotherapy for breast cancer: a randomized trial’. Breast, 21(1), p46–9. Available at: https://pubmed.ncbi.nlm.nih.gov/21865044/

3. Athas, W., Adams-Cameron, M., Hunt, W., Amir-Fazli, A., R Key, C. (2000), ‘Travel distance to radiation therapy and receipt of radiotherapy following breast-conserving surgery’, J Natl Cancer Inst, 92(3), p269–71. Available at: https://pubmed.ncbi.nlm.nih.gov/10655446/

4. Bargello-Rocha, J.E., et al. (2017), ‘The impact of the use of intraoperative radiotherapy on costs, travel time and distance for women with breast cancer in the Mexico City Metropolitan Area’, J Surg Oncol, 116(6), p683–689. Available at: https://pubmed.ncbi.nlm.nih.gov/28608393/.

5. Braun, V., Clarke, V., (2006). ‘Using thematic analysis in psychology.’ Qual Res Psychol, 3(2), pp77–101. Available at: 10.1191/1478088706qp063oa.

6. Byrne, D., (2021). ‘A worked example of Braun and Clarke’s approach to reflexive thematic analysis’, Quality & Quantity, 56, pp1391–1412.

7. Celaya, M., R Rees, J., J Gibson, J., L Riddle, B., Robert Greenberg, E. (2006), ‘Travel distance and season of diagnosis affect treatment choices for women with early-stage breast cancer in a predominantly rural population (United States)’, Cancer Causes Control, 17(6), p851–6. Available at: https://pubmed.ncbi.nlm.nih.gov/16783613/

8. Chierchini, S., Ingrosso, G., Saldi, S., Stracci, F., Aristei, C., (2019), ‘Physician and patient barriers to radiotherapy service access: treatment referral implications.’ Cancer Manag Res, 11, p8829–8833. Available at: https://www.ncbi.nlm.nih.gov/pmc/articles/PMC6789154/.

9. Christian, N., Sperk, E., Welzel, G., Abo-Madyan, Y., Kraus-Tiefenbacher, U., Keller, A., Sütterlin, M., and Wenz, F. (2012). ‘ TARGIT-E(lderly)—Prospective phase II study of intraoperative radiotherapy (TARGIT-IORT) in elderly patients with small breast cancer’, BMC Cancer, 12(171). Available at: https://www.ncbi.nlm.nih.gov/pmc/articles/PMC3519765/.

10. Coombs, N., Coombs, J., Vaidya, U., Singer, J., Bulsara, M., Tobias, J., Wenz, F., Joseph, D., Brown, D., Rainsbury, R., Davidson, D., Adamson, D., Massarut, S., Morgan, D., Potyka, I., Corica, T., Falzon, M., Williams, M., Baum, M., Vaidya, J. (2016), ‘Environmental and social benefits of the targeted intraoperative radiotherapy for breast cancer: data from UK TARGIT-A trial centres and two UK NHS hospitals offering TARGIT IORT’, BMJ open, 6, e010703. Available at: https://bmjopen.bmj.com/content/6/5/e010703

11. Corica, T., Saunders, Ch., Bulsara, M., Taylor, M., Joseph, D., Nowak, A. (2014), ‘Intraoperative radiotherapy for early breast cancer: do health professionals choose convenience or risk?’, Radiat Oncol, 25, p 9–33. Available at: https://pubmed.ncbi.nlm.nih.gov/24461031/

12. Corica, T., Nowak, A., Saunders, Ch., Bulsara, M., Taylor, M., Vaidya, J., Baum, M., Joseph, D. (2016). ‘Cosmesis and Breast-Related Quality of Life Outcomes After Intraoperative Radiation Therapy for Early Breast Cancer: A Substudy of the TARGIT-A Trial’ Radiation Oncology, 96(1), P55–64. Available at: https://www.redjournal.org/article/S0360-3016(16)30135-3/fulltext#%20

13. Corica, T., Nowak, A., Saunders, Ch., Bulsara, M., Taylor, M., Vaidya, J., Baum, M., Joseph, D., Williams, N., Keshtgar, M. (2018), ‘Cosmetic outcome as rated by patients, doctors, nurses and BCCT.core software assessed over 5 years in a subset of patients in the TARGIT-A Trial’, Radiation Oncology, 13(68). Available at: https://ro-journal.biomedcentral.com/articles/10.1186/s13014-018-0998-x

14. Corica, T., Saunders, Ch., Bulsara, M., Taylor, M., Joseph, D., Nowak, A. (2019), ‘Patient preferences for adjuvant radiotherapy in early breast cancer are strongly influenced by treatment received through random assignment’, Eur J Cancer Care (Engl), 28(2), e12985. Available at: https://pubmed.ncbi.nlm.nih.gov/30637839/

15. General Medical Council (GMC)., (2020). ‘Guidance on professional standards and ethics for doctors - Decision making and consent’. Available at: https://www.gmc-uk.org/-/media/documents/gmc-guidance-for-doctors---decision-making-and-consent-english_pdf-84191055.pdf

16. Goyal, S.H., Chandwani, S.H., G. Haffty, B., Demissie, K. (2015). ‘Effect of Travel Distance and Time to Radiotherapy on Likelihood of Receiving Mastectomy’, Breast Oncology, 22, p1095–1101. Available at: https://link.springer.com/article/10.1245/s10434-014-4093-8

17. Guidolin, K., Lock, M., Brackstone, M. (2018). ‘Patient-perceived barriers to radiation therapy for breast cancer’, Can J Surg, 61(2), p121–143. Available at: https://www.ncbi.nlm.nih.gov/pmc/articles/PMC5866151/.

18. Haviland, J.S., Hopwood, P., Mills, J., Sydenham, M., Bliss, J.M., and Yarnold, J.R. (2021). ‘Do patient-reported outcome measures agree with clinical and photographic assessments of normal tissue effects after breast radiotherapy? The experience of the standardisation of breast radiotherapy (START) trials in early breast cancer’, Clinical Oncology, 28(6), pp. 345–353. Available at: https://www.sciencedirect.com/science/article/pii/S0936655516000376.

19. Hickok, J.T., Morrow, G.R., Roscoe, J.A., Mustian, K. and Okunieff, P. (2019). ‘Occurrence, severity, and longitudinal course of twelve common symptoms in 1129 consecutive patients during radiotherapy for cancer’, Joint Pain Symptom Management, 30(5), pp. 433–442. Available at: https://www.sciencedirect.com/science/article/pii/S0885392405004744.

20. Keshtgar, M., Bulsara, M., Williams, N., Saunders, Ch., Flyger, H., Cardoso, J., Corica, T., Bentzon, N., Michalopoulos, N., Joseph, D. (2013), ‘Objective assessment of cosmetic outcome after targeted intraoperative radiotherapy in breast cancer: results from a randomised controlled trial’. Breast Cancer Research and Treatment, 140, pp.519–525. Available at: https://link.springer.com/article/10.1007/s10549-013-2641-8#citeas

21. Llewellyn, A., Howard, C. and McCabe, C. (2019), ‘An exploration of the experiences of women treated with radiotherapy for breast cancer: Learning from recent and historical cohorts to identify enduring needs’, European Journal of Oncology Nursing, 39, pp.47–54. Available at: https://www.sciencedirect.com/science/article/abs/pii/S146238891930002X.

22. Long, L.E. (2001), ‘Being informed: undergoing radiation therapy’, Cancer Nursing, 24(6), pp. 463–468. Available at: https://www.sciencedirect.com/science/article/pii/S107881742030198X#bib11.

23. Murchison, S., Soo, J., Inglesew, P.A., Hamilton, S. (2020). ‘Breast cancer patients’ perceptions of adjuvant radiotherapy: an assessment of pre-treatment knowledge and informational needs’, Journal of Cancer Education, 35(4), pp.661–668. Available at: https://link.springer.com/article/10.1007/s13187-019-01507-4.

24. National Institute of Health and Care Excellence (NICE), (2018). ‘Intrabeam radiotherapy system for adjuvant treatment of early breast cancer’. Available at: https://www.nice.org.uk/guidance/ta501/chapter/4-Committee-discussion.

25. Nattinger, A., T Kneusel, R., G Hoffmann, R., A Gilligan, M. (2001), ‘Relationship of distance from a radiotherapy facility and initial breast cancer treatment’, J Natl Cancer Inst, 93(17), p1344–6. Available at: https://pubmed.ncbi.nlm.nih.gov/11535710/

26. Pickles, K., Hersch, J., Nickel, B., Vaidya, J., McCaffery, K., Barratt, K. (2022), ‘Effects of awareness of breast cancer overdiagnosis among women with screen-detected or incidentally found breast cancer: a qualitative interview study’, BMJ open, 12, e061211. Available at: https://bmjopen.bmj.com/content/12/6/e061211

27. Probst, H., Rosbottom, K., Crank, H., Stanton, A. and Reed, H. (2021). ‘The patient experience of radiotherapy for breast cancer: A qualitative investigation as part of the SuPPORT 4 All study’, Radiography, 27(2), pp.352–359. Available at: https://www.sciencedirect.com/science/article/pii/S107881742030198X#bib14.

28. Schnur, J.B., Ouellette, S.C., Bovbjerg, D.H., and Montgomery. G.H., (2009). ‘Breast cancer patients’ experience of external-beam radiotherapy’, Qualitative Health Research, 19(5), pp.668–676. Available at: https://journals.sagepub.com/doi/abs/10.1177/1049732309334097.

29. Schnur, J.B., Ouellette, S.C., DiLorenzo, T.A., Green, S., and Montgomery, G.H. (2011). ‘A qualitative analysis of acute skin toxicity among breast cancer radiotherapy patients’, Psycho-Oncology, 20(3), pp.260–268. Available at: https://onlinelibrary.wiley.com/doi/abs/10.1002/pon.1734.

30. Shaverdian, N., Wang, X., Hegde, J.V., Aledia, C., Weidhaas, J.B., Steinberg, M.L., and McCloskey, A. (2018). ‘The patient’s perspective on breast radiotherapy: Initial fears and expectations versus reality’, Cancer, 124(8), pp.1673–1681. Available at: https://acsjournals.onlinelibrary.wiley.com/doi/full/10.1002/cncr.31159.

31. Sosin, M., Sen Gupta, S., Wang, J.S., Costellic, C.D., Gulla, A., Bartholomew, A.J., O’Neill, S.C., Hechenbleikner, E.M., Collins, B.T., Langan, R.C., Willey, S.C., and Tousimis, E.A. (2018), ‘A Prospective Analysis of Quality of Life and Toxicity Outcomes in Treating Early Breast Cancer With Breast Conservation Therapy and Intraoperative Radiation Therapy, Frontiers in Oncology, 8(545). Available at: https://www.frontiersin.org/articles/10.3389/fonc.2018.00545/full.

32. Stanton, A.L., Krishnan, L., Collins. M.A., (2001). ‘Form or function? Part 1. Subjective cosmetic and functional correlates of quality of life in women treated with breast-conserving surgical procedures and radiotherapy’, Cancer, 91(12), pp.2273–2281. Available at: https://acsjournals.onlinelibrary.wiley.com/doi/10.1002/1097-0142%2820010615%2991%3A12%3C2273%3A%3AAID-CNCR1258%3E3.0.CO%3B2-1.

33. Tang, T., Cohan, T., Beattie, G., Cureton, E., Svahn, J., Lyon, L., Kelly, J., Shim, V. (2021), ‘Patients Older 65 Years With Early Breast Cancer Prefer Intraoperative Radiation as a Locoregional Treatment Choice’, Ann Surg Oncol, 28(9), p 5158–5163. Available at: https://pubmed.ncbi.nlm.nih.gov/33751295/

34. Tong A, Sainsbury P, Craig J. ‘Consolidated criteria for reporting qualitative research (COREQ): a 32- item checklist for interviews and focus groups.’ Int J Qual Health Care. 2007;19(6):349–57.

35. Vaidya J, Vyas J, Chinoy R, Merchant N, Sharma O, Mivra I. ‘Multicentricity of breast cancer: Whole-organ analysis and clinical implications.’ British journal of cancer 1996;74(5):820–24.

36. Vaidya JS, Baum M, Tobias JS, D’Souza DP, Naidu SV, Morgan S, Metaxas M, Harte KJ, Sliski AP, Thomson E. ‘Targeted intra-operative radiotherapy (TARGIT): an innovative method of treatment for early breast cancer.’ Annals of oncology : official journal of the European Society for Medical Oncology / ESMO 2001;12(8):1075–80.

37. Vaidya, J.S., Joseph, D., Tobias, J.S., Bulsara, M., Wenz, F., Saunders, Ch., Alvarado, M., Flyger, H., Massarut, S., Eiermann, W., Keshtgar, M., Dewar, J., Kraus-Tiefenbacher, U., Süverlin, M., Esserman, L., MR Holtveg, H., Roncadin, M., Pigorsch, S., Metaxas, M., Falzon, M., Mavhews, A., Corica, T., R. Williams, N., Baum, M. (2010), ‘Targeted intraoperative radiotherapy versus whole breast radiotherapy for breast cancer (TARGIT-A trial): an international, prospective, randomised, non-inferiority phase 3 trial’, The Lancet, Fast track, 376(9735), p91–102. Available at: https://www.thelancet.com/journals/lancet/article/PIIS0140673610608379/fulltext#%20

38. Vaidya JS, Bulsara M, Baum M, Wenz F, Massarut S, Pigorsch S, Alvarado M, Douek M, Saunders C, Flyger H, Eiermann W, Brew-Graves C, Williams N, Potyka I, Roberts N, Bernstein M, Brown D, Sperk E, Laws S, Suverlin M, Corica T, Lundgren S, Holmes D, Vinante L, Bozza F, Pazos M, Le Blanc-Onfroy M, Gruber G, Polkowski W, Dedes KJ, Niewald M, Blohmer J, McCready D, Hoefer R, Kelemen P, Petralia G, Falzon M, Joseph D, Tobias JS. ‘Long term survival and local control outcomes from single dose targeted intraoperative radiotherapy during lumpectomy (TARGIT-IORT) for early breast cancer: TARGIT-A randomised clinical trial.’ BMJ 2020;370:m2836. Available at: hvps://www.bmj.com/content/bmj/370/bmj.m2836.full.pdf

39. Vaidya JS, Bulsara M, Baum M, Wenz F, Massarut S, Pigorsch S, Alvarado M, Douek M, Saunders C, Flyger H, Eiermann W, Brew-Graves C, Williams NR, Potyka I, Roberts N, Bernstein M, Brown D, Sperk E, Laws S, Suverlin M, Corica T, Lundgren S, Holmes D, Vinante L, Bozza F, Pazos M, Blanc-Onfroy ML, Gruber G, Polkowski W, Dedes KJ, Niewald M, Blohmer J, McReady D, Hoefer R, Kelemen P, Petralia G, Falzon M, Joseph D, Tobias JS. ‘New clinical and biological insights from the international TARGIT-A randomised trial of targeted intraoperative radiotherapy during lumpectomy for breast cancer.’ British journal of cancer 2021;125(3):380–89. Available at: hvps://www.nature.com/arwcles/s41416-021-01440-8.pdf

40. Vaidya, J.S., Vaidya, U.J., Baum, M., Bulsara K., M., Joseph, D., Tobias, J.S. on behalf of TARGIT-IORT Global Authors. (2022), ‘Global adoption of single-shot targeted intraoperative radiotherapy (TARGIT- IORT) for breast cancer—berer for patients, berer for healthcare systems’, Radiation Oncology, 12. Available at: https://www.frontiersin.org/journals/oncology/articles/10.3389/fonc.2022.786515/full

41. Welzel, G., Boch, A., Hofmann, F., Kraus-Tiefenbacher, U., Gerhardt, A., Suetterlin, M. and Wenz, F. (2013). ‘Radiation-related quality of life parameters after targeted intraoperative radiotherapy versus whole breast radiotherapy in patients with breast cancer: results from the randomized phase III trial TARGIT-A’, Radiation Oncology, 8(9). Available at: https://ro-journal.biomedcentral.com/articles/10.1186/1748-717X-8-9#citeas.

42. Williams, F. and Jeanetta, S.C. (2016). ‘Lived experiences of breast cancer survivors after diagnosis, treatment and beyond: qualitative study’, Health Expectations, 19(3), pp.631–642. Available at: https://onlinelibrary.wiley.com/doi/full/10.1111/hex.12372.

